# Comparison of disability level between Early and Late Onset Parkinson’s Disease using WHODAS 2

**DOI:** 10.1101/2023.08.31.23294866

**Authors:** Isaíra Almeida Pereira da Silva Nascimento, Kátia Cirilo Costa Nobrega, Bruno Rafael Antunes Souza, Isabela Carneiro Barone, Giovanna Checchio, Vitória Pereira Ponciano, Clara Greif Cerveira de Paula, Arieni Nunes Possani, Natália Cardoso Penha, André Frazão Helene, Antonio Carlos Roque, Rodolfo Savica, Maria Elisa Pimentel Piemonte

**Affiliations:** Department of Physical Therapy, Speech Therapy, and Occupational Therapy, Faculty of Medical Science, University of São Paulo, São Paulo, Brazil; Department of Neuroscience and Behavior, Institute of Psychology, University of São Paulo, Brazil; Department of Physiology, Institute of Biosciences, University of São Paulo, São Paulo, Brazil; Department of Physics, School of Philosophy, Sciences, and Letters of Ribeirão Preto, University of São Paulo, Ribeirão Preto, Brazil; Department of Neurology, Mayo Clinic, Rochester, MN, USA

**Keywords:** Parkinson’s disease, early onset, disability, cognition, WHODAS 2

## Abstract

**Background:** Parkinson’s disease (PD) is a degenerative neurological disorder that usually affects people over the age of 60. However, 10-20% of patients have an early onset of PD (EOPD).

**Objectives:** To compare disability levels according to the World Health Organization Disability Assessment Schedule 2.0 (WHODAS-2) between people with EOPD and those with late-onset PD (LOPD).

**Methods:** We conducted a cross-sectional study with 95 EOPD patients (mean-age 44.51±4.63, H&Y 1.93±0.93) and 255 LOPD patients (mean-age 63.01±7.99, H&Y 2.02±0.95). Demographic information, clinical characteristics, cognitive evaluation by Telephone-Montreal-Cognitive-Assessment (T-MoCA), functionality self-evaluation by WHODAS-2 and the Unified-Parkinson’s-Disease-Rating-Scale (MDS-UPDRS), parts I and II, were documented for each patient by an individual remote interview.

**Results:** Analysis showed a statistically significant difference between EOPD and LOPD in the cognition and activities of daily-living-related to work/school domains of WHODAS-2. T-MoCA scores confirmed more impaired cognition capacity in LOPD. The two groups had no significant differences in levodopa daily dosage, Hoehn and Yahr (H&Y) stages, disease time duration, and MDS-UPDRS I and II scores.

**Conclusion:** People living with EOPD face similar disability levels as those with LOPD, except for cognition, where LOPD patients exhibited higher levels of disability than EOPD and for work activities where the EOPD exhibited higher levels of disability than LODP. These results highlight the challenges faced by people with EOPD in interacting with society and living with the disease for a longer time. The WHODAS-2 can be a useful tool to assess disability and tailor interventions for people with PD of different age groups.

## 1. Introduction

Parkinson’s disease (PD) is the second most common neurodegenerative disorder after Alzheimer’s dementia [1]. The disease onset is typically around 65 to 70 years of age [2], and the higher prevalence (2%) is found in people over the age of 70, defined as Late Onset of PD (LOPD) [3]. However, Early Onset of PD (EOPD), defined as PD with age at onset after 21 years but before 50 years [4], also be observed, with an incidence between 0.29 and 3.3 per 100,000 persons-years [5,6].

Given the effect of PD symptoms earlier in life, the interaction of symptoms with potentially more active roles in society, and the longer time-life living with the disease, people with EOPD face more challenges than those with LOPD. Rigidity, and dystonia have been found as more frequently presenting symptoms in EOPD [7,8]. In addition, the severity of the postural instability after a longer disease duration is higher in people with EOPD than in LOPD [9]. Interestingly, regarding non-motor symptoms, the rate of depression was reported higher in patients with EOPD than in patients with LOPD [10], as well as worse emotional well-being and poorer quality of life, independent of depression status. [11].

Besides the motor and non-motor aspects, people with EOPD face different social challenges than LOPD. Unemployed men with EOPD are twice more than what observed in the general population [12], and early retirement is also higher for people with EOPD [13]. Finally, EOPD may present a challenge to relationships. Couples with EOPD reported higher marital discord scores than those with LOPD [14]. Sexual dysfunction has been reported as more prevalent in EOPD than in the general population [15].

The interaction among the physical, mental, and social aspects may be behind the higher prevalence of suicidal ideas in people with EOPD than in LOPD [16]. Despite these specific challenges, there is no standardized pharmacological and non-pharmacological therapeutic approach for EOPD. Better awareness about the EOPD disability will undoubtedly improve the management and care of these patients [17].

When it comes to controlling the severity of Parkinson’s Disease, objective measures based on motor tests have proven to be quite effective. However, it’s crucial to understand that the impact of the disease cannot always be directly linked to motor impairments alone, even though there is a clear relationship [18]. There seems to be a notable gap between how healthcare providers and patients view the health condition linked to Parkinson’s disease, according to a recent report by movement disorder specialists from around the world [19]. The current preconized care model centered on the patient [20] emphasizes the significance of self-perception in assessing disability levels as the primary indicator of health conditions. This approach recognizes the significance of the patient’s own experience and perspective as the primary indicator of their health condition.

The World Health Organization Disability Assessment Schedule 2.0 (WHODAS-2) was developed to assess disability based on the International Classification of Functioning, Disability, and Health (ICF) [21]. According to ICF, disability is described as”a difficulty in functioning at the body, person, or societal levels, in one or more life domains, as experienced by an individual with a health condition in interaction with contextual factors’’ [22]. Although other tools have traditionally been used to measure disability, such as the Indexes of activities of daily living and quality of life, none of them has been developed based on the ICF biopsychosocial conceptual model [21].

A seminal study to assess the WHODAS-2 conceptual model and metric properties in a set of chronic and prevalent clinical conditions accounting for a broad scope of disability included people living with early, intermediate, and advanced PD stages among the 1,119 participants recruited in 7 European centers. The results showed very high reliability, good ability to discriminate among known groups, and adequate capacity to detect change over time, confirming that WHODAS-2 is adequate to evaluate disability in patients with chronic conditions, which may help to eliminate barriers to developing policies, giving excellent evidence of these populations’ need [21].

The results from a cross-sectional analytical study examining the metric properties WHODAS-2 short version, which included 168 PD patients with 69.6 mean age, demonstrated their suitable metric properties [23].

Regarding the complex interaction among the motor, mental and social aspects associated with disability level in people living with EOPD and being WHODAS-2, a common, international, and interdisciplinary instrument based on the biopsychosocial model, the present study aimed to investigate the disability level in people living with EOPD in comparison with people living with LOPD.

## 2. Materials and Methods

### 2.1. Study Design and Participants

A cross-sectional study of 350 people with idiopathic PD was recruited for this study. The people were divided into the EOPD group (age of onset between 21 and 50 years, n = 95, mean age 44.51±4.63, mean H&Y stage 1.93±0.93) and LOPD group (age of onset >50 years, n = 255, mean age 63.01±7.99, mean H&Y stage 2.02±0.95) (Figure 1). The eligibility criteria were (a) a diagnosis of idiopathic PD according to the diagnostic criteria of the UK Parkinson’s Disease Society Brain Bank [24], confirmed by a neurologist, and (b) ≥ 21 years old [4], `and (c) being treated for PD in the 6 months preceding the commencement of the study and (d) have access to telephone or internet and agree to participate in the study`. The non-eligibility criteria were (a) the presence of neurological disorders other than PD and (b) the presence of dementia, speech, and hearing disorders since interviews were conducted by phone calls or phone messages.

**FIGURE 1.**
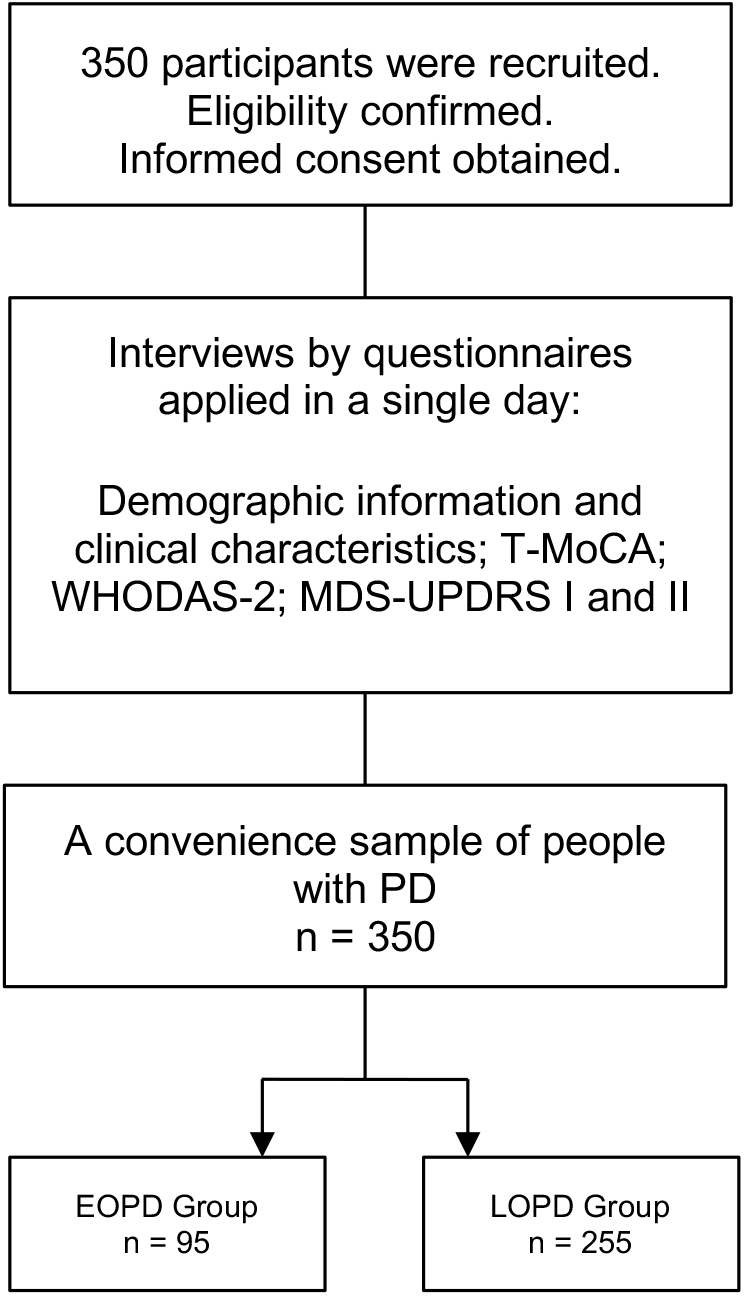
The schematic study designs. T-MoCA, Telephone-Montreal Cognitive Assessment; WHODAS-2, World Health Organization Disability Assessment Schedule 2; MDS-UPDRS, Movement Disorder Society - Unified Parkinson’s Disease Rating Scale; EOPD, Early-onset Parkinson’s disease; LOPD, Late-onset Parkinson’s disease.

To assess the cognitive status, we considered the ability of the participants to properly answer the first section of the study questionnaire about personal and socioeconomic information as clinical evidence of the minimal cognitive capacity to self-evaluate their health condition.

### 2.2. Recruitment

Participants were recruited from health units that offer care for PD. At first, through calls and/or telephone messages, we identified their eligibility. Subsequently, information about the study procedures was passed on, and they were invited to consent to participate.

This study was approved by the proper Ethics Committee and conducted in accordance with the Helsinki Declaration.

### 2.3. Study Procedures

The participants were asked to indicate the best day and time for the remote interview and if a family member could help them to answer the questions. Then, the researchers applied the questionnaire in a single day, which included: 1- general information (i.e., age, sex/gender, race), 2- socioeconomic status, 3- information associated with PD, 4- information dosage of medication [25], 5- self-evaluation on functionality by World Health Organization Disability Assessment Schedule - WHODAS-2 [23], 6- self-evaluation on”non-motor aspects of daily life experiences” by MDS-UPDRS (Part I): [26], 7- self-evaluation on”motor experiences daily living” by - MDS-UPDRS (Part II) [26] and 8- evaluation of the global cognitive capacity by T-MoCA [27].

### 2.4. Instruments

#### 2.4.1. WHODAS-2 (World Health Organization Disability Assessment Schedule 2.0)

It is a practical, generic assessment instrument that can measure health and disability at the population level or clinical practice based on the difficulties presented in the last 30 days. There are several different versions of WHODAS-2, which differ in length and intended mode of administration. This study used the full version of 36 items through an interview.

WHODAS-2 captures the level of functioning in six domains of life (3): Domain 1: Cognition – understanding and communicating; Domain 2: Mobility – moving and getting around; Domain 3: Self-care – attending to one’s hygiene, dressing, eating, and staying alone; Domain 4: Getting along – interacting with other people; Domain 5: Life activities – domestic responsibilities, leisure, work, and school; Domain 6: Participation – joining in community activities, participating in society.

For all six domains, WHODAS-2 provides a profile and a summary measure of functioning and disability that is reliable and applicable across cultures in all adult populations. Each question is scored between 1 and 5. The higher the score, the worse the functionality of people.

#### 2.4.2. T-MoCA (Telephone - Montreal Cognitive Assessment)

The T-MoCA is an adapted version of the MoCA 30 administered over the telephone with changes in the score. We used the MoCA items that did not require the use of a pencil and paper, or a visual stimulus were used for the T-MoCA, which has a maximum score of 22.

### 2.5 Statistical Analysis

Descriptive statistical analysis was used for the demographic and clinical data of the participants. Considering the normal age distribution, test T was used to compare the two groups (EOPD and LOPD).

The Kruskal-Wallis ANOVA was used for disease stages comparison (1, 2, and 3 stages according to H&Y classification). In addition, the Mann-Whitney U Test with continuity correction was used for group comparison (EOPD and LOPD).

Finally, the Spearman Rank Order Correlation was used to test the correlation between MDS-UPDRS I-II and WHODAS-2 scores and between T-MoCA and WHODAS-2 domain cognitive scores. Differences were considered statistically significant when the p-value <0.05. The statistical analyses were performed using Statistica Version 13 (TIBCO Software Inc. USA).

## 3. Results

The demographic and clinical participants’ features are demonstrated in Table 1. As expected, most of them are males, with a significant statistical difference in age between the groups. There were no statistically significant differences in levodopa daily dosage, H&Y stages, disease time duration, and MDS-UPDRS I and II scores.

**TABLE 1.**
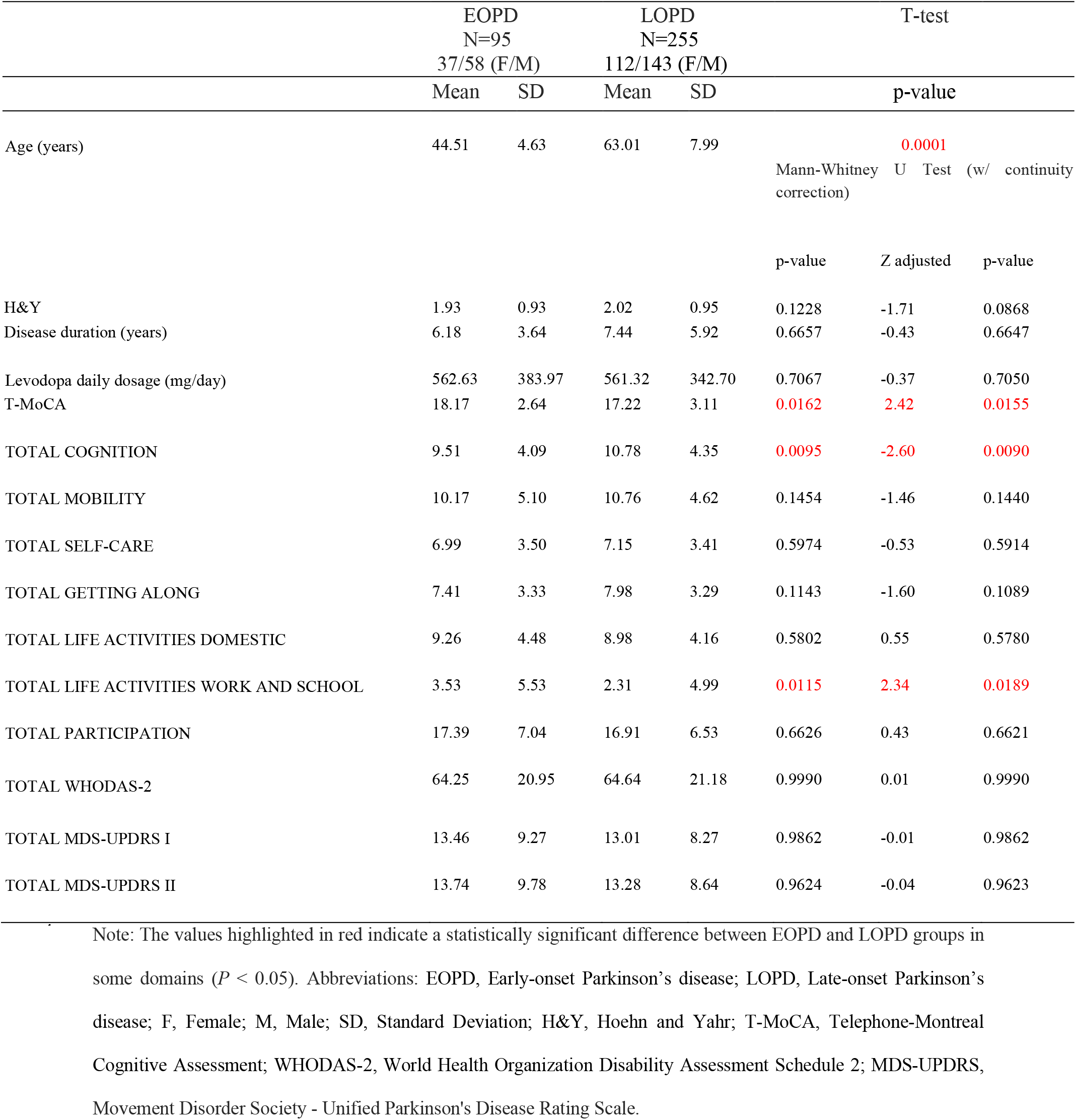
Demographic and clinical characteristics of people with idiopathic PD.

Regarding the disease evolution, there was a statistically significant difference in WHODAS-2 total scores and MDS-UPDRS I and II scores between the intermediate disease stage (H&Y 3) and early disease stages (H&Y 1 and 2), independent of the group (p-value=.0000001; .00007; .0000001 respectively), whereas there was not a statistically significant difference in T-MoCA scores among the H&Y stages. Additionally, there was a positive significant correlation between UPDRS I and II and WHODAS-2 scores (R=.61, p-value=.00001 and R=.65, p-value=.00001, respectively).

Regarding the disease onset, there was a statistically significant difference between EOPD and LOPD groups in two domains of WHODAS-2 only: cognition and activities of daily living (ADL) associated with work/school. For cognition, the participants of LOPD group reported higher disability levels than EOPD (Figure 2). Furthermore, the analysis of T-MoCA scores reinforced this finding: participants from LOPD showed statistically significantly lower scores than participants from EOPD, which reflects a worse cognitive global performance (Figure 3). Lastly, WHODAS-2 domain cognitive and T-MoCA scores were inversely and significantly correlated (R= - .304, p-value=.0000001).

**FIGURE 2.**
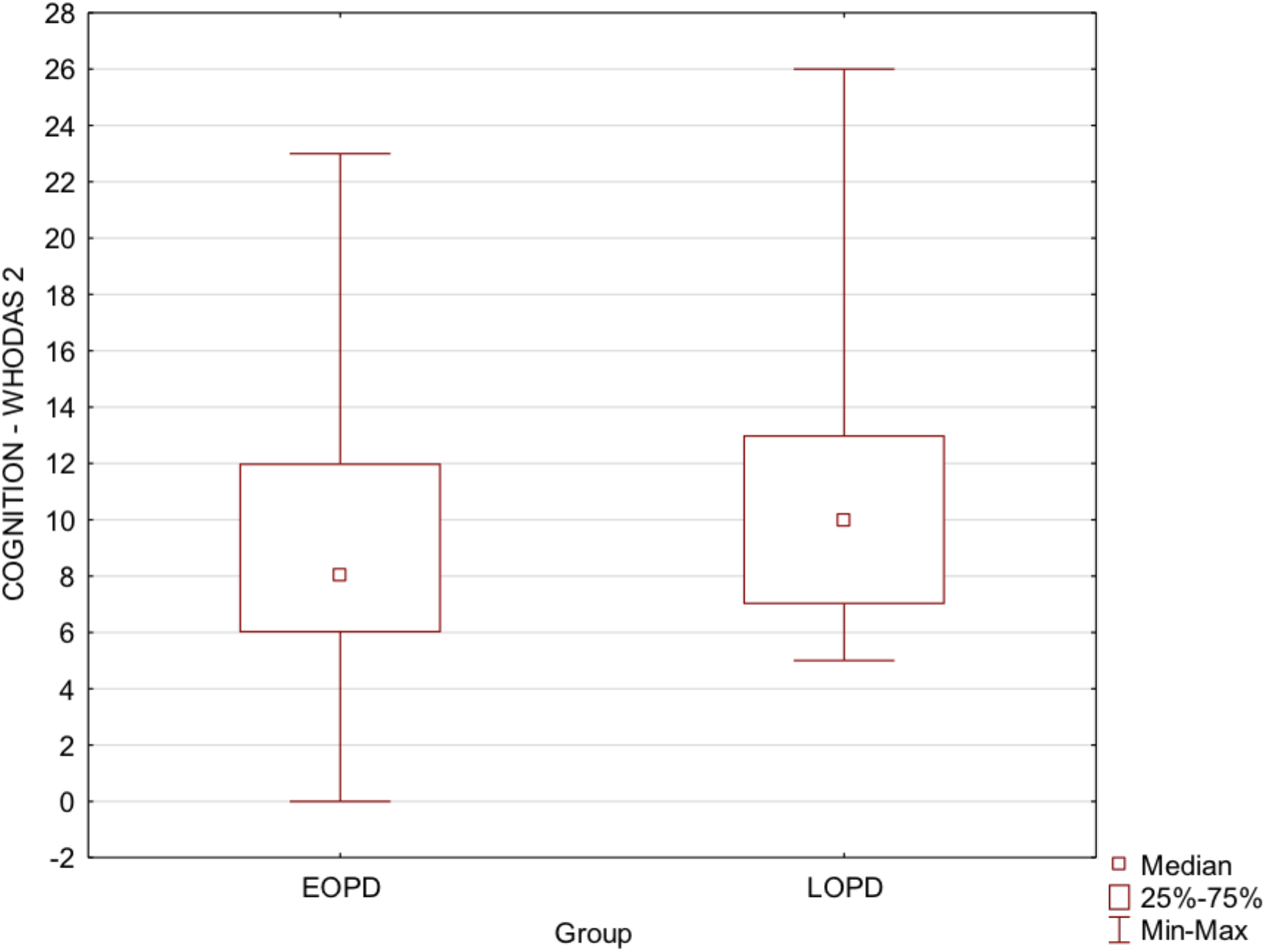
Box plots show EOPD and LOPD group Cognition-WHODAS 2 scores. Squares represent means, lines represent medians, and boxes show the interquartile range. Maximum and minimum scores are indicated by the upper and lower markers, respectively. EOPD, Early-onset Parkinson’s disease; LOPD, Late-onset Parkinson’s disease; WHODAS-2, World Health Organization Disability Assessment Schedule 2.

**FIGURE 3.**
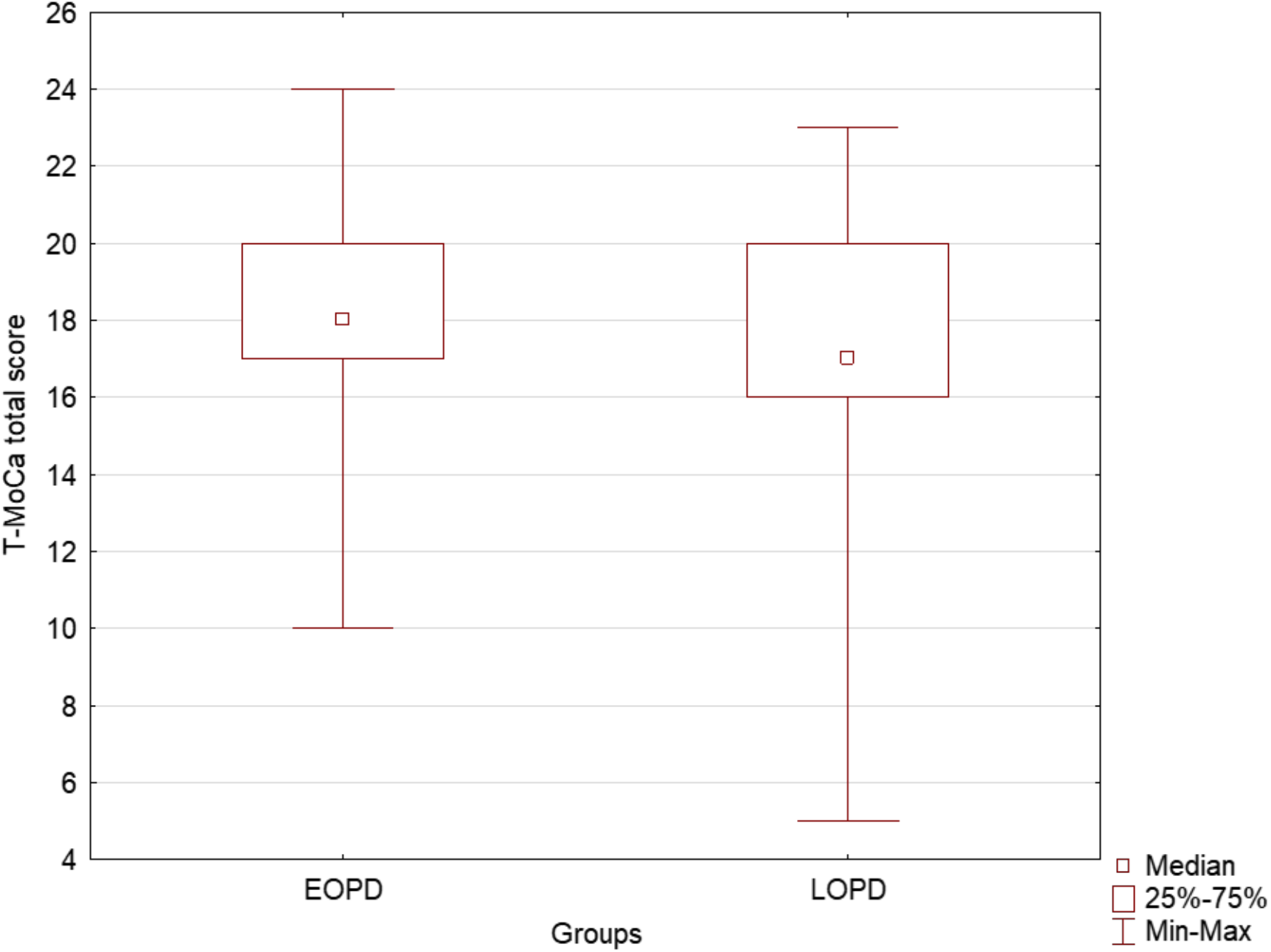
Box plots show EOPD and LOPD group T-MoCA total scores. Squares represent means, lines represent medians, and boxes show the interquartile range. Maximum and minimum scores are indicated by the upper and lower markers, respectively. EOPD, Early-onset Parkinson’s disease; LOPD, Late-onset Parkinson’s disease; T-MoCA, Telephone-Montreal Cognitive Assessment.

In contrast, for working ADL, the participants from EOPD group reported higher disability levels than LOPD. Additional analysis showed that 5% of participants from the LOPD group reported being”unemployed” against 11% from the EOPD group, 36% to be”employed” against 41% from the EOPD group, and 50% were”retired” against 28% from the EOPD group.

## 4. Discussion

Overcoming a neurodegenerative, progressive, and incurable disease such as PD is a complex challenge in any life cycle. Supposedly, managing the motor and non-motor symptoms and their social consequences is harder for younger people in the more active phase of life. In fact, the results from the current study showed a worse disability level and higher unemployment rate in people living with EOPD. However, the self-perception of disability levels related to mobility, self-care, getting along, life activities, and participation were similar for people living with EOPD and LOPD. In contrast, self-perception cognitive disability level was higher for people with LOPD than EOPD. Several considerations can be made based on our results.

First, the WHODAS-2 was able to detect differences between clinical-severity groups: participants classified as intermediate disease stage (H&Y 3) reported worse disability scores than those with early disease stages (H&Y 1 and 2). Additionally, the disability levels according to WHODAS-2 were correlated with motor and non-motor aspects of daily living experiences according to MDS-UPDRS, considered golden standing for PD evaluation [26]. The WHODAS-2, designed to cover disability, measures the restrictions on daily life activities and social participation [21], whereas MDS-UPDRS I and II address the impact of non-motor and motor symptoms on daily life activities [26]. The moderate magnitude of the associations among the two instruments reflects how the WHODAS-2 and the MDS-UPDRS I and II measure different aspects of related concepts. Thus, WHODAS-2 may be used to complement the information obtained by MDS-UPDRS, offering a more comprehensive evaluation of the biopsychosocial impact of disease on patient functionality.

Second, despite some previous studies showing the slower disease evolution for early onset [28], our results showed a similar disability level according to WHODAS-2 and motor and non-motor aspects of daily living experiences according to MDS-UPDRS for both groups that reported the same disease duration. These findings show that people with early and late-onset PD experience similar disability levels in daily living after living with the disease for the same amount of time. The UPDRS Section II has been found to be a reliable tool for measuring disease progression, particularly in the early stages of the disease [29] and has stronger and more stable association with disease duration than other UPDRS sections [30]. Sections I and II of the UPDRS also have established clinically important differences that can be used to judge the effectiveness of interventions [31]. Therefore, self-reported experiences related to daily living can be considered a reliable tool to measure the severity and progression of PD. Based on this finding, it is plausible to suppose that the participants from EOPD could potentially experience higher disability levels when at the same age as LOPD participants. Further longitudinal studies are needed to investigate this issue. However, this reasonable supposition should be alert for specialized care improvement for people with EOPD.

Third, differences were found between the two groups for two WHODAS-2 domains: cognition and work/school activity. The disability level related to cognition was higher for LOPD participants. The decline in global cognitive capacity due to the aging process is well-known [32]. The interaction between neurodegeneration associated with PD and aging processes may explain why younger people living for a similar time with PD have a smaller cognitive disability. Results from T-MoCA confirm the more preserved cognitive global capacity for EOPD participants than LOPD. However, T-MoCA was not able to show the difference between the disease stages regardless of the onset. In contrast, WHODAS-2 domain cognitive showed significantly higher disability levels for participants in the intermediate PD stage (H&Y 3) in comparison to initial PD stages (H&Y 1 and 2). This finding suggests that even before objective tests can detect cognitive decline, they can impact cognitive functionality. Then, cognitive intervention should be considered priority care for people with LOPD and be started even before the detectable objective impairments.

Finally, the higher disability levels in work activities for EOPD than LOPD was unsurprising. A previous study comparing people with EOPD with LOPD showed that, among those who retired, 97% of the patients with EOPD retired early versus 73% of those with LOPD [14]. Other studies have found that 54% of people with EOPD retire early, and 94% are likely to give up work within ten years of disease onset [13]. In the present study, 28% of participants reported being retired, even having less than 60 years of age. Another previous study showed that men aged 55 to 64 living with PD were twice as likely to be unemployed than the general population [12]. In the present study, the percentage of unemployed was twice larger for EOPD than for LOPD participants (11% and 5%, respectively). Finally, 20% of people with EOPD reported being away from their jobs for health reasons. Further studies are needed to investigate the long-term socioeconomic impact of this population’s high unemployment level and early retirement. Thus, besides offering proper care to minimize the effects of motor and non-motor symptoms on work activities performance, it is crucial to improve the social support for this population as well as to promote a social policy to decrease the social stigma.

Our study has some limitations. The absence of a motor evaluation of the motor symptoms severely may be considered a limitation of the present study; therefore, we cannot necessarily explore and correlate motor function and self-perception of disability level in the two groups. However, the importance of our findings is still maintained since the self-perception of disability level is a relevant measure that can provide valuable insight into the personal experiences of those living with PD [30,31]. In addition, our study is based on a relatively small number of patients; therefore, a larger sample size is needed to confirm our findings. Also, socioeconomic, and social status may have a major role in our results; however, we do not have detailed data to potentially explore this additional important variable. Lastly, a potential recall bias can be evoked when the participants have been interviewed via phone, but it should be equally distributed among EOPD and LOPD; unfortunately, patients with cognitive decline may have more difficulties in the phone interview and we should be aware about this possible limitation as well.

Further studies are needed to investigate the association between disability level according to WHODAS-2 and PD motor symptoms severity.

## 5. Conclusion

In our study, we observed similar global disability levels between people living with EOPD and LOPD, according to WHODAS-2, except in the cognition domain that was more impaired in people living with LOPD than EOPD, and in activities related to work that was more impaired in people living EOPD than LOPD, findings that can be partially explained by the productive cycle of life.

## 6. Data Availability Statement

The raw data supporting the conclusions of this article will be made available by the authors, without undue reservation.

## 8. Author contributions

IN: Research Project (organization and data collection), Manuscript Preparation (writing of the first draft), Review and Critique. KN: Research Project (organization and data collection), Manuscript Preparation (writing of the first draft). BS: Research Project (organization and data collection). IB, GC, VP, CP, AP and NP: Research Project (data collection). AH, AR and RS: Manuscript Preparation (writing of the first draft), Review and Critique. MP: Research Project (conception and organization), Manuscript Preparation (writing of the first draft), Review and Critique. All authors contributed to the article and approved the submitted version.

## 9. Acknowledgment

This study was financed in part by the Coordenação de Aperfeiçoamento de Pessoal de Nível Superior - Brasil (CAPES) - Finance Code 001. This article was produced as part of the activities of FAPESP Research, Innovation, and Dissemination Center for Neuromathematics (grant #<u>2013/ 07699-0</u>, S.Paulo Research Foundation).

## 11. Publisher’s note

All claims expressed in this article are solely those of the authors and do not necessarily represent those of their affiliated organizations, or those of the publisher, the editors and the reviewers. Any product that may be evaluated in this article, or claim that may be made by its manufacturer, is not guaranteed or endorsed by the publisher.

